# Rationale and design of the brain magnetic resonance imaging protocol for FutureMS: a longitudinal multi-centre study of newly diagnosed patients with relapsing-remitting multiple sclerosis in Scotland

**DOI:** 10.1101/2021.03.10.21253264

**Authors:** Rozanna Meijboom, Stewart J Wiseman, Elizabeth N York, Mark E Bastin, Maria del C Valdés Hernández, Michael J Thrippleton, Daisy Mollison, Nicole White, Agniete Kampaite, Koy Chong Ng Kee Kwong, David Rodriguez Gonzalez, Dominic Job, Christine Weaver, Patrick K A Kearns, Pete Connick, Siddharthan Chandran, Adam D Waldman

## Abstract

**Introduction:** Multiple sclerosis (MS) is a chronic neuroinflammatory and neurodegenerative disease. MS prevalence varies geographically and is notably high in Scotland. Disease trajectory varies significantly between individuals and the causes for this are largely unclear. Biomarkers predictive of disease course are urgently needed to allow improved stratification for current disease modifying therapies and future targeted treatments aimed at neuroprotection and remyelination. MRI can detect disease activity and underlying damage non-invasively *in vivo* at the micro and macrostructural level. FutureMS is a prospective Scottish longitudinal multi-centre cohort study, which focuses on deeply phenotyping patients with recently diagnosed relapsing-remitting MS (RRMS). Neuroimaging is a central component of the study and provides two main primary endpoints for disease activity and neurodegeneration. FutureMS aims to reduce uncertainty around disease course and allow for targeted treatment in RRMS by exploring the role of conventional and advanced MRI measures as biomarkers of disease severity and progression. This paper provides an overview of MRI data acquisition, management and processing in FutureMS.

**Methods and analysis:** MRI is acquired at baseline (N=431) and 1-year follow-up, in Dundee, Glasgow and Edinburgh (3T Siemens) and in Aberdeen (3T Philips), and managed and processed in Edinburgh. The core structural MRI protocol comprises T1-weighted, T2-weighted, FLAIR and proton density images. Primary imaging outcome measures are new/enlarging white matter lesions (WML) and reduction in brain volume over one year. Secondary imaging outcome measures comprise WML volume as an additional quantitative structural MRI measure, rim lesions on susceptibility-weighted imaging, and microstructural MRI measures, including diffusion tensor imaging and neurite orientation dispersion and density imaging metrics, relaxometry, magnetisation transfer (MT) ratio, MT saturation and derived g-ratio measures.

**Ethics and dissemination:** The study received ethical approval from the South East Scotland Research Ethics Committee 02 (reference 15/SS/0233). Results will be made available to study participants and funders.

**STRENGTHS AND LIMITATIONS OF THE STUDY:**

- Brain imaging acquired within six months after relapsing-remitting multiple sclerosis (RRMS) diagnosis allows for detection of abnormalities at a very early stage of disease.
- Longitudinal structural and microstructural brain MRI are integrated with multi-modal clinical, blood biomarker, genetic and retinal imaging data to allow comprehensive evaluation of early RRMS stratifiers.
- MR protocol optimization was implemented mid-study to improved image quality and protocol harmonization, resulting in some between-subject protocol variation.
- Multiplatform MRI acquisition required to allow access for a large MS population across Scotland introduces inevitable inter-site data variance.
- Contrast enhanced MRI was not acquired, limiting imaging evaluation of active inflammatory lesions.

## 1. INTRODUCTION

### Multiple sclerosis

Multiple sclerosis (MS) is a chronic debilitating disease of the central nervous system (CNS), for which a cure is not yet available. Pathology occurs in both white (WM) and grey matter (GM) in the brain and spinal cord and is characterised by inflammation-induced demyelination and neurodegeneration [1,2]. Worldwide there are over two million cases of MS [3]. Scotland has a notably high prevalence of the disease with 290 cases per 100,000 population [4–6]. People with MS experience a wide range of symptoms, including mobility and vision problems, cognitive impairment and fatigue [7]; the severity of which varies markedly between individuals. In 85-90% of cases, MS starts with a relapsing and remitting disease course (RRMS), which in later stages generally becomes progressive (secondary progressive MS; SPMS), and the remainder have a progressive course from onset (PPMS) [8,9]. The disease trajectory also varies significantly between individuals, the causes for which are largely unclear. Establishing early biomarkers predictive of disease course is highly important as it may allow appropriately targeted disease-modifying therapy (DMT).

### FutureMS

FutureMS (https://future-ms.org/) is a large (N=440) longitudinal multi-centre observational cohort study in Scotland aiming to develop predictive tools for disease progression and markers of disease severity in a deeply phenotyped early-stage RRMS cohort. A detailed cohort description is available in Kearns et al. 2021 [10]. Brain MR imaging, focusing on both structural and microstructural techniques, is a core feature of FutureMS and is importantly being studied in the context of the whole disease with participants undergoing extensive neurological, quality of life, cognitive, retinal imaging, blood biomarker and genomic assessments at each study visit. MRI-based biomarkers in MS are of great value, as MRI can be used to study CNS damage in vivo and non-invasively, offering potential as a predictor of future disability. Particularly, within the framework of a well-powered longitudinal study, changes in individual trajectory of MR measures that occur associated with treatment can be a powerful real-world marker of DMT efficacy.

### Structural MRI in MS

Conventional MR imaging plays an essential role in faster MS diagnosis [11] as well as MS research, particularly as DMT trial end-point [12]. Characteristic MS abnormalities seen on conventional MRI include periventricular, callosal, juxtacortical and infratentorial WM hyperintense lesions (WML) on T2-weighted (T2W) images; the central vein sign within WML on T2 images [13]; hypointensities (‘black holes’) on T1-weighted (T1W) images; and brain atrophy. Cortical brain lesions are difficult to detect on 1.5T and 3T MRI [11,12]. WML accrual is used as indicator of interval disease activity; and T1W hypointensities and gadolinium-enhanced lesions are used as respectively indicators of irreversible damage and active inflammation [11,12,14]. Clinical measures of disease severity/progression only show some association with brain abnormalities on conventional MRI [15], which is thought to be due to limitations in both clinical disability and imaging measures [16,17]. The number and location (i.e. infratentorial) of WML, can be predictive of conversion from clinically isolated syndrome to MS and accumulation of disability [18,19]. Similarly, enhancing WML and T1W ‘black holes’ have been associated with progression of disability [20–22]. In addition, although still an area of research, there is a growing body of evidence indicating that chronic active lesions (or rim/smouldering lesions) characterised by paramagnetic rims can be identified using susceptibility weighted imaging (SWI), and are associated with increased disability [23]. Spinal cord WML and atrophy are also commonly present, where damage confers a disproportionate risk of disability given the anatomical eloquence [24].

Processing techniques applied to conventional imaging allow for assessment of neurodegeneration through quantitative volumetric measures of whole-brain (GM and WM) atrophy. Whole-brain atrophy is present in early-stage RRMS [18,19], progresses over time and affects multiple brain regions [25]. It is associated with and predictive of clinical disability [16,18,26,27]. Although GM atrophy appears to be more closely associated with clinical progression [28,29] and seems to decrease more extensively over time compared with WM atrophy [30–32]. Overall their contribution to clinical disability remains largely unclear [33], and atrophy must be considered a downstream and non-specific indicator of neurodegeneration. Nonetheless, measures of brain volume are the most established imaging marker of neurodegeneration in MS, and have been proposed as the only measure currently sufficiently validated and reliable for use as a study endpoint [33].

### Microstructural MRI in MS

Macrostructural imaging methods are widely used but provide indirect markers of the underlying pathophysiological features and leave a great deal of disease progression unaccounted for [16]. Quantitative MRI techniques that provide more specific markers of brain integrity on the microstructural level are becoming more established for studying demyelination and neurodegeneration in MS and may better account for clinical consequences.

Magnetisation transfer (MT) imaging provides an indirect means of detecting protons bound to semi-solid macromolecules, e.g. myelin, that are not visible on conventional sequences, via the exchange of magnetisation with the directly detectable protons of free water molecules. The most widely used MT measure is the MT ratio (MTR), which is the fractional signal reduction caused by exchange of magnetisation between these free and bound protons [34,35]. The MTR is affected by demyelination and axonal integrity [34,36,37] and thus reflects microstructural WM abnormalities. Previous studies have shown associations between MTR and clinical disability in MS [12,38–41], indicating it may have potential as a biomarker. MTR signal is, however, also affected by biophysical parameters (notably T1 relaxation time and B1 inhomogeneities), compromising its specificity to tissue microstructure [42]. MT saturation (MTsat) imaging corrects for these parameters, and provides increased contrast between WM and GM, and may therefore provide a better biomarker of microstructural WM changes than MTR [42,43]. It has not been widely applied to MS, but associations with cognition and disability have been reported [42,44].

Quantitative multi-shell diffusion MRI (dMRI) provides additional markers of brain microstructure based on tissue water diffusion characteristics. Diffusion tensor imaging (DTI) models diffusion-dependent signal based on anisotropic water molecule displacement due to spatially ordered brain microarchitecture, which is particularly prominent within WM. A change in the WM microstructure will lead to a change of water molecule displacement [45]. DTI parameters, comprising fractional anisotropy (FA), mean diffusivity (MD), axial (λ_AX_) and radial (λ_RD_) diffusivity, are sensitive to changes in the microstructure that are relevant to MS pathology [45]. λ_RD_ is thought to be sensitive to demyelination [46] whereas axonal loss is mostly reflected in changes in λ_AX_ [47]. These metrics can be measured globally or regionally in tissue, but can also be combined with tractography methods [48] to assess WM tract-specific microstructural damage. In addition, multi-shell dMRI allows for neurite orientation dispersion and density imaging (NODDI) analysis, which enables more precise characterisation of WM microstructure, i.e. neurite (axon and dendrite) density, and dispersion of neurite orientation [49]. Previous pathological and MRI studies have shown that neurite density is affected in MS [50,51].

Combining MTI and dMRI allows calculation of an MRI-derived aggregate g-ratio. The g-ratio is the ratio between the inner (i.e. axon) and outer (i.e. axon and myelin) diameter of the WM fibre and reflects myelin thickness relative to the axon radius [52–54]. Preliminary studies in small subject groups have observed g-ratio abnormalities in MS, suggestive of a thinner myelin sheath, in accordance with known pathological changes in this disease [53,55,56].

Relaxometry maps based on transverse (T2) or longitudinal (T1) relaxation times are also used to study underlying tissue changes. Studies suggest that quantification of T1 and combined T1 and T2 measures can be correlated with myelin content in tissue [57], and that quantitative T2 mapping of WML yields additional information related to clinical disability [58].

### MRI in FutureMS

FutureMS incorporates a comprehensive MRI protocol, including visual and quantitative assessment of WML, GM and WM volumes, dMRI and MTI metrics, as well as g-ratio and relaxometry measures. These structural (conventional) imaging and quantitative microstructural metrics will be explored longitudinally with physical, cognitive and other quality of life features, blood biomarkers, genetics and retinal imaging, allowing for studying and developing predictive tools of disease progression and markers of disease severity in RRMS.

### Study aim

The aim of this paper is to provide a rationale and transparent overview of MRI acquisition and processing in FutureMS, including detailed descriptions of the MRI protocol, MRI data management and MRI processing pipelines.

## 2. METHODS

### 2.1 Participants

FutureMS [10] recruited patients with a recent diagnosis of RRMS (<6 months) [11] from five neurology hubs in Scotland: Edinburgh, Glasgow, Dundee, Aberdeen and Inverness. Further inclusion criteria were: aged 18 years or older, and the capacity to provide informed consent. Exclusion criteria were intake of DMTs prescribed prior to baseline assessment, participation in a clinical trial prior to baseline assessment and contraindications for MRI. Each participant received an MRI examination at baseline and 1-year follow-up, as well as a full neurological assessment, cognitive testing, and blood marker and genetic testing. Recruitment for FutureMS completed in March 2019, with a total of 440 participants (N=431 for MRI) included for baseline assessment. Nine participants did not undergo MRI mainly due to fulfilling MRI exclusion criteria. One-year follow-up was completed with 392 (N=386 for MRI) participants having returned for a follow-up visit. See Table 1 for demographics. Further follow-up visits will take place at 5-years and 10-years after baseline.

**Table 1.** Baseline demographics for MRI study participants

|  | N | Gender<br>(F/M) | Mean age<br>in years<br>(SD) | Mean<br>EDSS<br>(SD) |
| --- | --- | --- | --- | --- |
| RRMS | 431 | 317/114 | 38 (10.3) | 2.5<br>(1.3) |

F=female, M=male, SD=standard deviation, EDSS=expanded disability status scale, RRMS=relapsing-remitting multiple sclerosis

All patients were given a patient information sheet, had the study explained to them and gave written informed consent before study entry. The study received ethical approval from the South East Scotland Research Ethics Committee 02 under reference 15/SS/0233 and is being conducted in accordance with the Declaration of Helsinki and ICH guidelines on good clinical practice. All imaging data and additional clinical data were anonymised with unique study identifiers.

### 2.2 MRI acquisition protocol

All FutureMS participants received an MRI examination consisting of structural (conventional) MRI sequences. Additionally, a selection of participants underwent sub-study protocols including SWI (N=0 at baseline, N=44 at 1-year follow-up), and the microstructural imaging techniques multi-shell dMRI and MTI (for both: N=78 at baseline and N=67 at 1-year follow-up).

#### 2.2.1 Core study sequences: structural MRI

Structural MRI was acquired at four sites (Table 2, 3). The Glasgow and Dundee study sites used a Siemens Prisma 3T system and Aberdeen used a Philips Achieva 3T system. Edinburgh employed two different systems for the study. Participants included in the study from the start in May 2016 up to and including October 2017 have been imaged on a Siemens Verio 3T system (upgraded to Skyra Fit in July 2018; Site 1) and have received their follow-up MRI on the same system. Participants included in the study as of November 2017 until the end of recruitment in March 2019 have been imaged on a Siemens Prisma 3T system (Site 2) and have received their follow-up MRI on this system. Inverness study site participants were imaged at one of the other four sites.

**Table 2.** Equipment manufacturers across the sites participating in Future MS

| Site | Edinburgh (Site 1) | Edinburgh (Site 2) | Glasgow | Dundee | Aberdeen |
| --- | --- | --- | --- | --- | --- |
| **Manufacturer** | Siemens | Siemens | Siemens | Siemens | Phillips |
| **Model** | Verio/Skyra | Prisma | Prisma | Prisma | Achieva |
| **Head coil** | 12 ch | 32 ch | 20 ch | 20 ch | 32 ch |
| **Participants scanned at w0** | 98 | 88 | 161 | 46 | 36 |
| **Started scanning** | May 2016 | November 2017 | November 2016 | December 2016 | March 2017 |

**Table 3.**
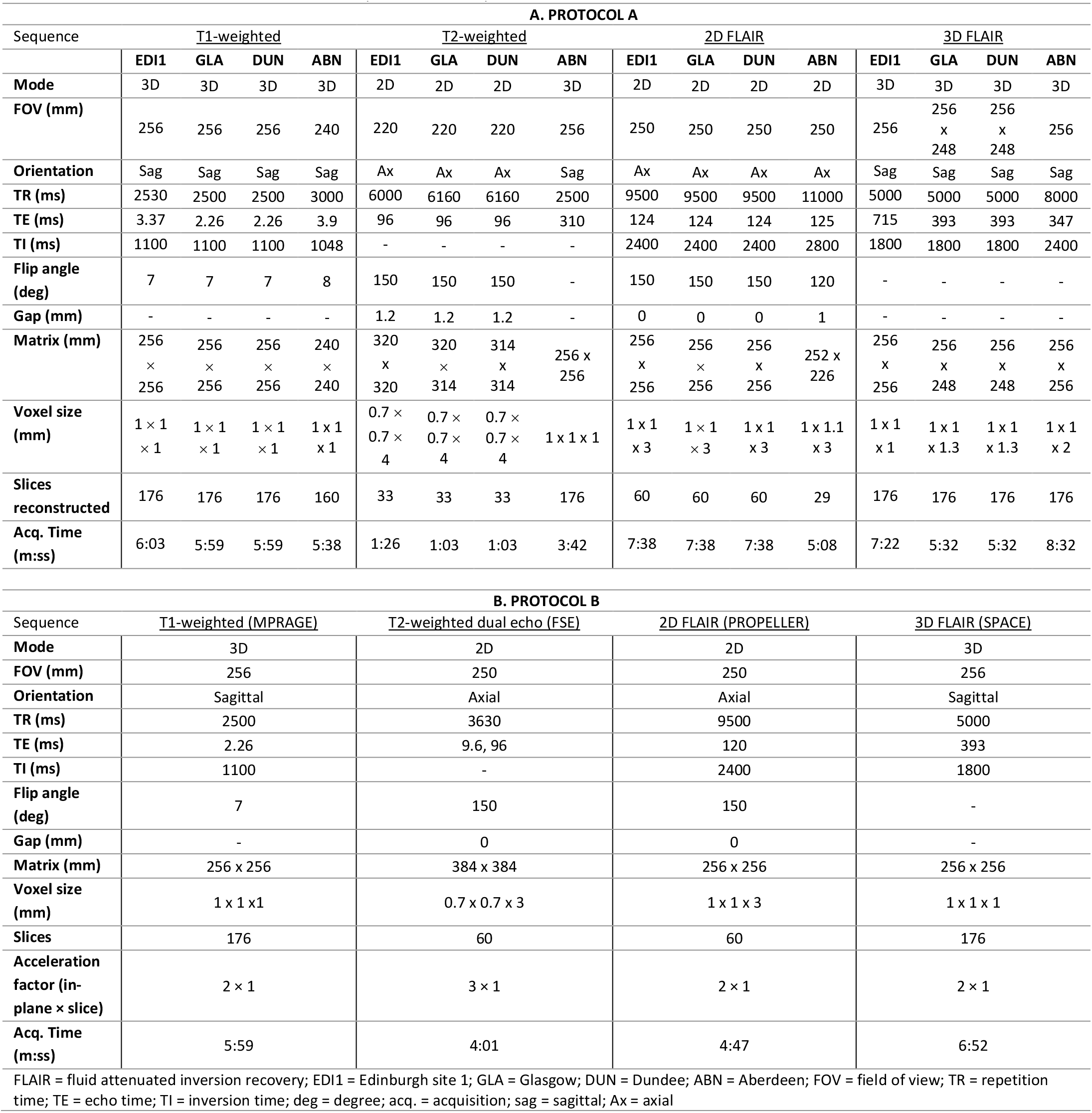
Future MS conventional MRI parameters for protocol A and B

In November 2017, the conventional MRI protocol was updated to increase between-site comparability and to facilitate improved image analysis. All participants who had been imaged before the update underwent protocol A (Table 3A), including their follow-up MRI. All patients scanned after the update were imaged with protocol B (Table 3B) at all visits. Protocol A (Table 3A) included an axial T2W, volumetric 3D T1W and two fluid attenuated inversion recovery- (FLAIR) weighted sequences: a 2D version with 3mm thick slices and no slice gap and a 3D version with 1 × 1 × 1 mm isotropic resolution. In accordance with the STRIVE guidelines [59], FLAIR was used to visualise WML. By definition, the imaging parameters of the FLAIR sequence were selected to supress fluid signals (specifically cerebrospinal fluid (CSF)) allowing other (pathological) fluids and tissues to become conspicuous. Protocol B (Table 3B) included 3D T1W, dual echo and 2D and 3D FLAIR sequences. A dual echo sequence provided T2W and proton density (PD) images, which allowed for T2 mapping for inflammation assessment [60] and for more accurate extraction of the intracranial volume (ICV). Additionally, for protocol B, parameters of all sequences were matched as closely as possible across sites to obtain maximum comparability. An overview of structural images is provided in Figure 1.

**Figure 1.**
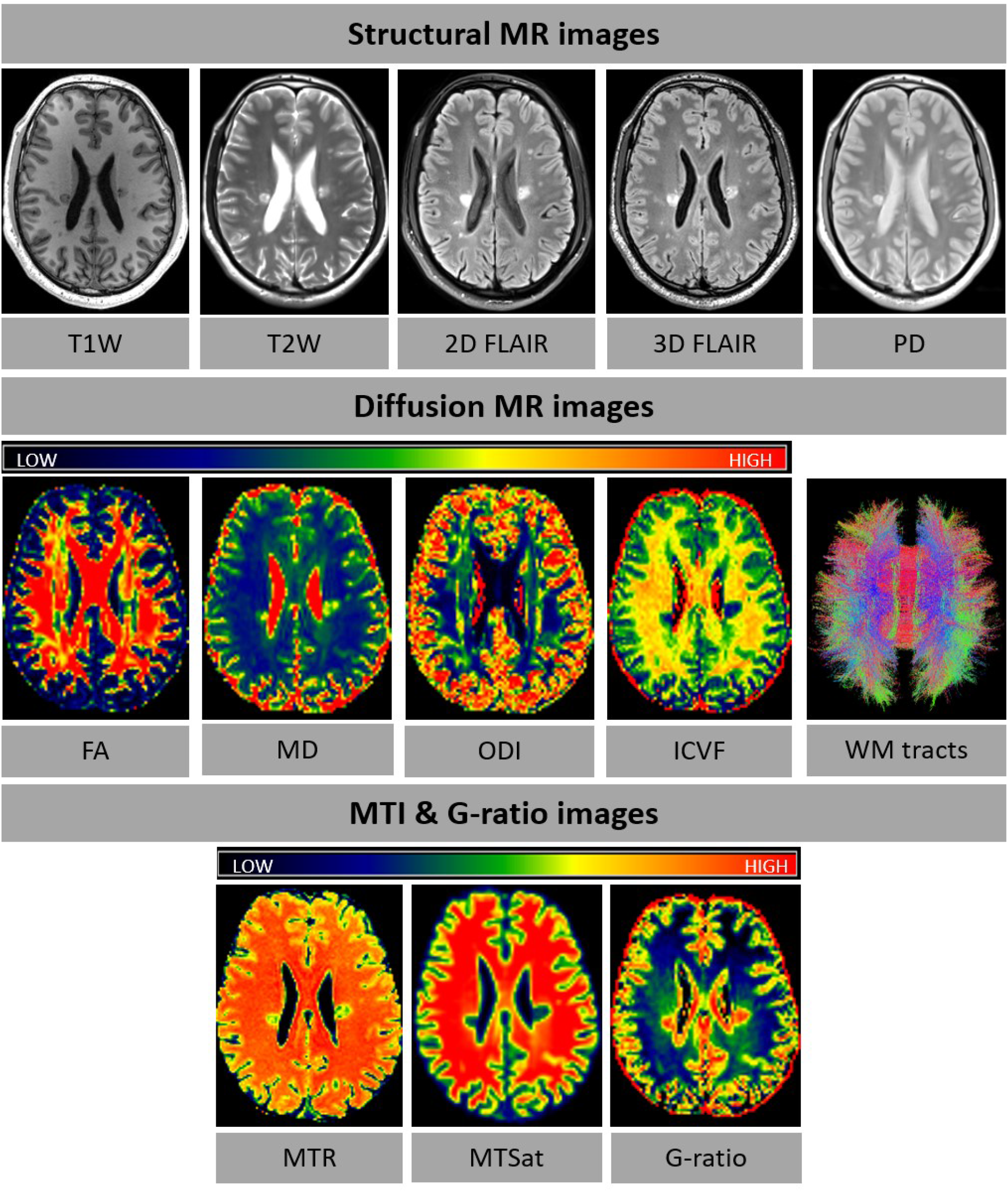
Examples of FutureMS MR images for structural MRI and microstructural MRI (diffusion MRI, MTI and g-ratio). dMRI, MT and G-ratio images are colour-coded according to the colour spectrum shown above the respective images. In the WM tract image, red is for right-left, blue for dorsal-ventral, and green for anterior-posterior tracts. Abbreviations: T1W=T1-weighted, T2W=T2-weighted, PD=proton density, FA=fractional anisotropy, MD=mean diffusivity, ODI=orientation dispersion index, ICVF=intracellular volume fraction, WM=white matter, MTI=magnetisation transfer imaging, MTR=magnetisation transfer ratio, MTSat=magnetisation transfer saturation.

#### 2.2.2 Sub-study sequences: SWI, dMRI and MTI

The sub-study sequences were imaged at a selection of study sites. The SWI sequence (Table 4) was acquired in Edinburgh (Site 2), Glasgow and Dundee, and implemented to investigate chronic inflammation in WML, as reflected by rim lesions. The sequence was set up as described in Sati et al. (2017) [61]. The SWI sub-study was added at 1-year follow-up visits only. Multi-shell dMRI [62] and MTI (Table 4; Figure 1) were acquired in Edinburgh (Site 2) and implemented to study myelin damage at a microstructural level. Optimised water diffusion-encoding magnetic field gradient vectors were generated as described in Caruyer et al. (2013) [62]. The sub-study sequences were combined with protocol B into a single examination.

**Table 4.** FutureMS MRI sub-study parameters

| Sequence | <u>MT on/MT off/T1W</u><br>Multi-echo spoiled<br>gradient echo | <u>dMRI</u> | <u>SWI</u> |
| --- | --- | --- | --- |
| **Mode** | 3D | 2D | 3D |
| **FOV (mm)** | 224 (SI) x 241 (AP) | 256 | 250 x 218.75 |
| **Directions** | - | 151 | - |
| **b-value (no. of directions)**<br>**(s/mm<sup>2</sup>)** | - | 0<sup>rev</sup> (3), 0 (14), 200 (3),<br>500 (6), 1000 (64), 2000<br>(64) | - |
| **Orientation** | Sagittal | Axial | Sagittal |
| **TR (ms)** | 30/30/15 | 4300 | 64 |
| **TE (ms)** | 1.54, 4.55, 8.49 | 74 | 35 |
| **Flip angle (deg)** | 5/5/18 | - | 10 |
| **Gap (mm)** | 0 | 0 | 0 |
| **Matrix (mm)** | 160 x 172 | 128 x 128 | 384 x 336 |
| **Voxel size (mm)** | 1.4 x 1.4 x 1.4 | 2 x 2 x 2 | 0.65 x 0.65 x 0.65 |
| **Slices** | 128 | 74 | 288 |
| **Acceleration factor (in-**<br>**plane x slice)** | 2 x 1 | 2 x 2 | - |
| **Acq. Time (m:ss)** | 6:14/6:14/3:08 | 11:12 | 7:08 |

MRI = magnetic resonance imaging; FOV = field of view; TR = repetition time; TE = echo time; deg = degree; acq. = acquisition; rev = reverse phase-encode direction; MT = magnetization transfer; T1W = T1-weighted; dMRI = diffusion MRI; SWI = susceptibility weighted imaging

### 2.3 Data storage

All data were anonymised before they were transferred to the imaging research team. The recommendations of the Brain Imaging Data Structure (BIDS; v1.0.1) (http://bids.neuroimaging.io/)[63] were followed for data storage.

### 2.4 Quality control

All raw imaging data were visually inspected for gross errors (e.g. ghosting and movement artefacts) [64] and data that were identified as inadequate were excluded from further processing.

### 2.5 Imaging outcomes

#### 2.5.1 Qualitative assessment: WML progression

WML progression was established as a binary outcome of the presence of new/enlarging lesions at 1-year follow-up, based primarily on the FLAIR volume sequence.

#### 2.5.2 Qualitative assessment: rim lesions on SWI

Presence of rim lesions was established for participants with SWI images acquired at 1-year follow-up. Rim lesions (Figure 2) were defined as hyperintense lesions on FLAIR which also have a hyperintense core surrounded by a hypointense rim on SWI.

**Figure 2.**
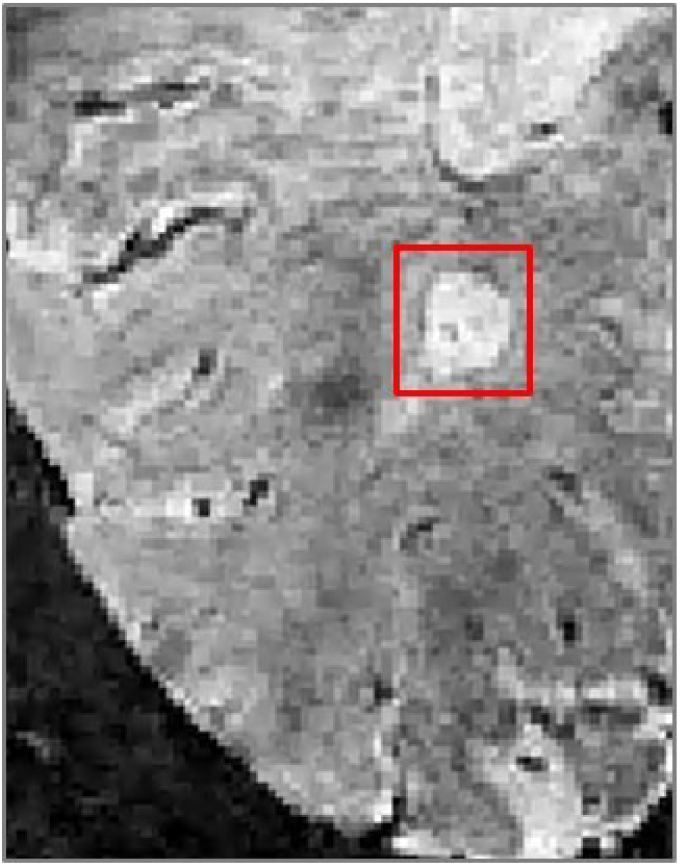
A paramagnetic hypointense rim lesion in relapsing-remitting multiple sclerosis identifying chronic inflammation, as visible on susceptibility weighted imaging.

#### 2.5.3 Quantitative assessment: macrostructural outcomes

Tissue volumes for total brain, regional and global cerebral and global cerebellar normal-appearing WM (NAWM), regional and global cerebral and global cerebellar cortical GM, subcortical GM (amygdala, thalamus, hippocampus, ventral diencephalon and basal ganglia), brainstem and WML were calculated in native space for baseline and follow-up (Table 5). Tissue volume change over one year was derived for each tissue type separately by subtracting baseline from follow-up volumes.

**Table 5.**
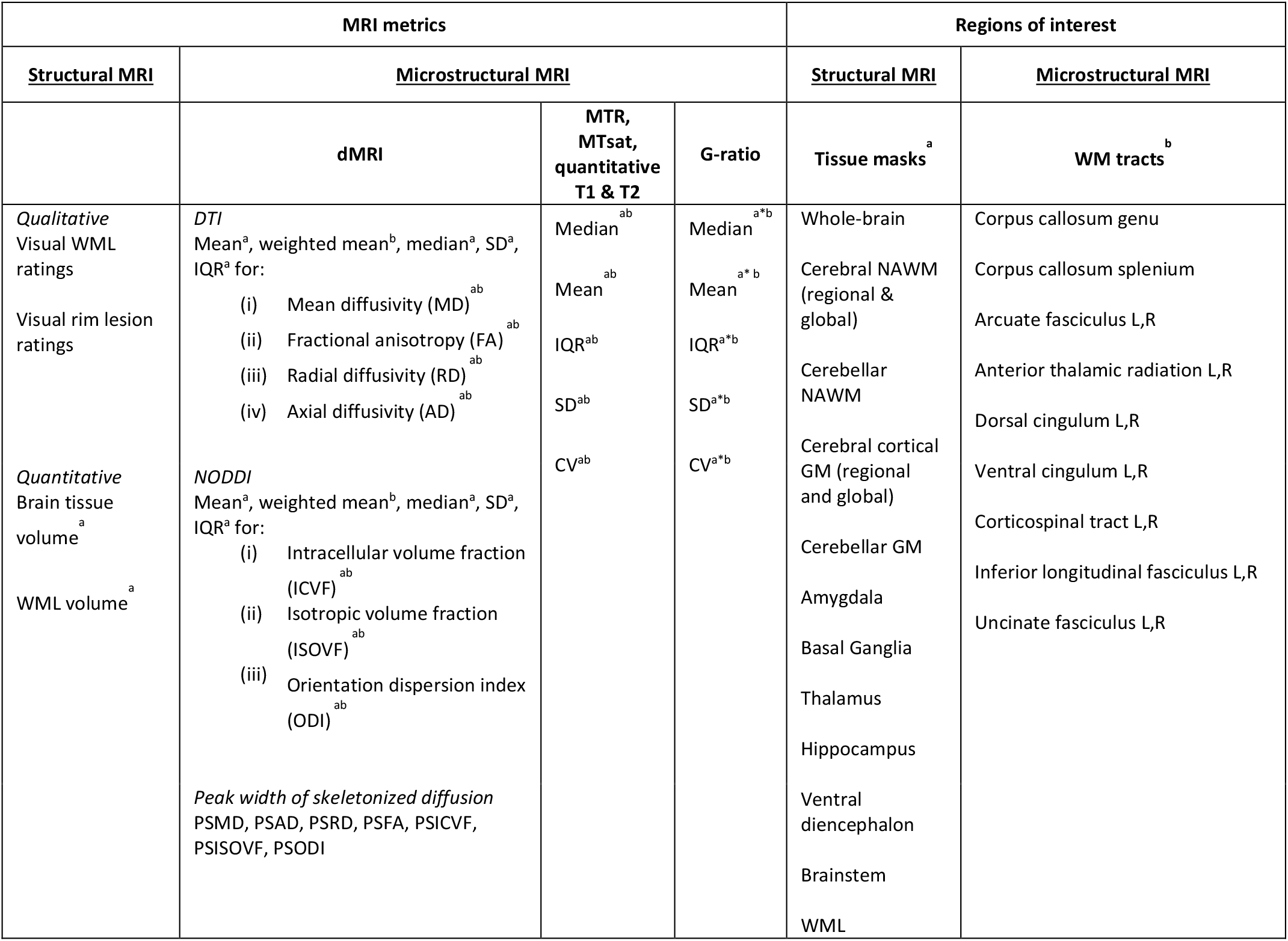
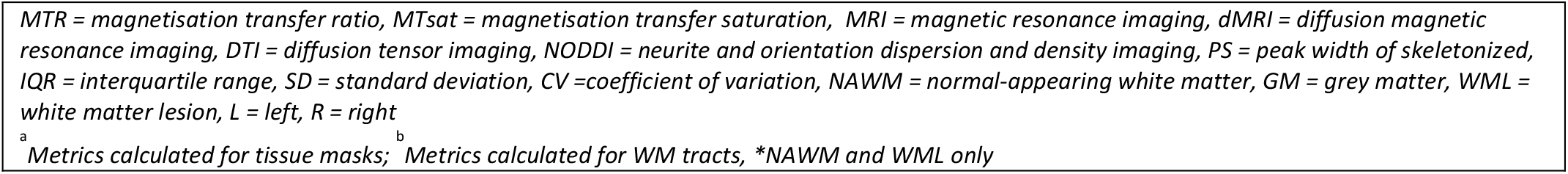
Overview of imaging variables and regions of interest

| MRI metrics |  |  |  | Regions of interest |  |
| --- | --- | --- | --- | --- | --- |
| Structural MRI | Microstructural MRI |  |  | Structural MRI | Microstructural MRI |
|  | dMRI | MTR, MTsat, quantitative T1 & T2 | G-ratio | Tissue masks<sup>a</sup> | WM tracts<sup>b</sup> |
| Qualitative Visual WML ratings | DTI<br>Mean<sup>a</sup>, weighted mean<sup>b</sup>, median<sup>a</sup>, SD<sup>a</sup>, IQR<sup>a</sup> for:<br>(i) Mean diffusivity (MD)<sup>ab</sup><br>(ii) Fractional anisotropy (FA)<sup>ab</sup><br>(iii) Radial diffusivity (RD)<sup>ab</sup><br>(iv) Axial diffusivity (AD)<sup>ab</sup> | Median<sup>ab</sup> | Median<sup>a*b</sup> | Whole-brain | Corpus callosum genu |
| Visual rim lesion ratings |  | Mean<sup>ab</sup> | Mean<sup>a*b</sup> | Cerebral NAWM (regional & global) | Corpus callosum splenium |
|  |  | IQR<sup>ab</sup> | IQR<sup>a*b</sup> | Cerebellar NAWM | Arcuate fasciculus L,R |
|  |  | SD<sup>ab</sup> | SD<sup>a*b</sup> | Cerebral cortical GM (regional and global) | Anterior thalamic radiation L,R |
| Quantitative Brain tissue volume<sup>a</sup> | NODDI<br>Mean<sup>a</sup>, weighted mean<sup>b</sup>, median<sup>a</sup>, SD<sup>a</sup>, IQR<sup>a</sup> for:<br>(i) Intracellular volume fraction (ICVF)<sup>ab</sup><br>(ii) Isotropic volume fraction (ISOVF)<sup>ab</sup><br>(iii) Orientation dispersion index (ODI)<sup>ab</sup> | CV<sup>ab</sup> | CV<sup>a*b</sup> | Cerebellar GM | Dorsal cingulum L,R |
| WML volume<sup>a</sup> |  |  |  | Amygdala | Ventral cingulum L,R |
|  |  |  |  | Basal Ganglia | Corticospinal tract L,R |
|  |  |  |  | Thalamus | Inferior longitudinal fasciculus L,R |
|  | Peak width of skeletonized diffusion<br>PSMD, PSAD, PSRD, PSFA, PSICVF, PSISOVF, PSODI |  |  | Hippocampus | Uncinate fasciculus L,R |
|  |  |  |  | Ventral diencephalon |  |
|  |  |  |  | Brainstem |  |
|  |  |  |  | WML |  |

MTR = magnetisation transfer ratio, MTsat = magnetisation transfer saturation, MRI = magnetic resonance imaging, dMRI = diffusion magnetic resonance imaging, DTI = diffusion tensor imaging, NODDI = neurite and orientation dispersion and density imaging, PS = peak width of skeletonized, IQR = interquartile range, SD = standard deviation, CV = coefficient of variation, NAWM = normal-appearing white matter, GM = grey matter, WML = white matter lesion, L = left, R = right
<sup>a</sup> Metrics calculated for tissue masks; <sup>b</sup> Metrics calculated for WM tracts, \*NAWM and WML only

#### 2.5.4 Quantitative assessment: microstructural outcomes

For dMRI, the mean for FA, MD, λ_AX_ and λ_RD,_ and intracellular volume fraction (ICVF), isotropic volume fraction (ISOVF) and orientation dispersion index (ODI) was determined for the white matter skeleton (PSMD, PSAD, PSRD PSFA, PSICVF, PSISOVF and PSODI; see below). Additionally, weighted means for these measures were determined within sixteen tracts of interest identified using quantitative tractography; and their mean, median, interquartile range (IQR) and standard deviation (SD) were determined within the above brain tissue compartments (total brain, WML, cerebral and cerebellar cortical NAWM, cerebral and cerebellar cortical GM and subcortical GM) at baseline and follow-up (Table 5). Similarly, mean, median, IQR, SD and coefficient of variation (CV) for MTR, MTsat, g-ratio, quantitative T1 and T2 were established within the same WM tracts and brain tissue compartments (only NAWM and WML for the g-ratio) at baseline and follow-up (Table 5).

### 2.6 Image processing

All DICOM images (http://dicom.nema.org) were converted to NIfTI-1 (http://nifti.-nimh.nih.gov/nifti-1) using dcm2niix (https://github.com/rordenlab/dcm2niix/releases), except for dMRI data, which were converted using the TractoR package (http://www.tractor-mri.org.uk) [65].

#### 2.6.1 Qualitative assessment: WML progression

Visual rating outcomes of WML progression were determined by an experienced neuroradiologist (DM). All standard structural sequences for baseline and follow up imaging were reviewed using the Carestream image viewing software (www.carestream.com), with a binary outcome of the presence of new/enlarging lesions at follow up based primarily on the FLAIR volume sequence. A random sample of 10% of studies were reviewed to assess intra- and inter-rater reproducibility, by the same observer following a delay of at least four weeks, and separately by a second experienced neuroradiologist (ADW).

#### 2.6.2 Qualitative assessment: rim lesions on SWI

Visual rating of rim lesions were assessed on SWI. Follow-up 2D FLAIR images were registered to SWI with a rigid body registration (degrees of freedom (DOF) = 6) using FSL FLIRT (FSL6.0.1; https://fsl.fmrib.ox.ac.uk/fsl) [66,67]. WML were identified on registered 2D FLAIR and assessed in all three anatomical planes on corresponding SWI images using ITK-SNAP3.8.0 [68]. Rim lesions were defined as lesions that are hyperintense on FLAIR, and are characterised by a hyperintense core partially or completely surrounded by a hypointense rim on SWI. Possible rim lesions were identified by a trained observer (KCNKK) and reviewed by a senior neuroradiologist (ADW). SWI images were also independently assessed for rim lesions by a second neuroradiologist (DM). Final rim lesion count for each subject was determined by consensus of all three raters. WML deemed too small to be reliably assessed for the presence of a rim were excluded. WML located near a high density of veins or considerable juxtacortical signal heterogeneity cannot be reliably evaluated and were therefore not considered for inclusion.

#### 2.6.3 Structural image processing: registration and brain extraction

At each time-point, T2W, PD and 2D FLAIR images were linearly registered to the T1W image with a rigid body transformation (6 DOF) using FSL FLIRT (FSL6.0.1) [66,67]. Brain extraction was performed on baseline scans only using FSL BET2 (FSL6.0.1) [69] with different settings for each acquisition protocol. Protocol A used T1W and T2W for brain extraction while protocol B used the PD volume. The resulting baseline ICV masks were visually checked and manually edited where required using ITK-SNAP3.8.0 [68]. The baseline edited ICV mask was then registered to the follow-up image space.

#### 2.6.4 Structural image processing: WML segmentation

WML segmentation was performed on the baseline FLAIR image using an adjusted method from Zhan et al. (2014) [70]. FLAIR hyperintense tissue voxels were identified by thresholding the raw brain image intensities to values higher than 1.69 times the standard deviation above the mean. This number was tested and optimized for the current study. A lesion distribution probabilistic template generated from a sample of 277 individuals with different degrees of WML, as per Chen et al. (2015) [71], was then applied to the thresholded image for excluding any hyperintense areas unlikely to reflect pathology (e.g. those produced by CSF flow artefacts around the third ventricle, in the WM tracts running perpendicular to the acquisition plane, near some sulci, and temporal poles). Further refinement of the resulting image was achieved by applying a Gaussian smoothing, followed by thresholding the image again to remove voxels with an intensity value z-score < 0.95 (z-scores were calculated in the raw FLAIR image) and by thresholding the then remaining voxel intensity values with threshold < 0.1. The resulting WML mask was binarised, checked and edited where necessary using ITK-SNAP [68]. For follow-up, the edited baseline WML masks were registered to follow-up space and re-edited to include any follow-up lesion changes.

#### 2.6.5 Structural image processing: Tissue segmentation

Tissue segmentation and brain parcellation was performed using FreeSurfer6.0 (http://surfer.nmr.mgh.harvard.edu/). For each wave separately (cross-sectional), tissue segmentation was performed on the T1W and T2W images, using the default parameters (including the Desikan-Kiliany atlas [72]) and the edited ICV as brain mask. The edited ICV masks were converted to FreeSurfer space and file format using mri_convert (https://surfer.nmr.mgh.harvard.edu/fswiki/mri_convert). All FreeSurfer tissue segmentations were visually assessed using an in-house snapshot software script. For incorrect segmentations, the appropriate files were corrected using FreeView2.0 after which FreeSurfer was rerun using the corrected files. Segmentations that remained incorrect after manual editing were discarded. For longitudinal analysis, FreeSurfer’s longitudinal processing stream [73] was applied to the cross-sectional data of all waves. The longitudinal processing stream reduces random variation to increase sensitivity for correct detection of changes over time. The above described cross-sectional data were combined to form an unbiased and subject-specific template with common information of the time points. This template was then used as a base for tissue segmentation for each time point separately (longitudinal). Visual checks and corrections of longitudinal output were performed as described above. Cross-sectional and longitudinal FreeSurfer tissue segmentations were converted to NIfTI format and native space using respectively Freesurfer’s mri_label2vol (https://surfer.nmr.mgh.harvard.edu/fswiki/mri_label2vol) and mri_convert. FreeSurfer output was corrected for WML load where appropriate, using fslmaths (FSL6.0.1), resulting in tissue masks as described in section 2.5.3. Tissue volumes, as well as WML and ICV volumes, were then extracted using (FSL6.0.1).

#### 2.6.6 dMRI processing: DTI and NODDI

FSL6.0.1 tools, including FSL topup and eddy [74], were used to extract the brain, remove bulk motion and geometric/eddy current induced distortions by registering all subsequent volumes to the first T2W echo-planar (EP) volume [66], estimate the water diffusion tensor and calculate parametric maps of MD, λ_AX_ and λ_RD_, and FA from its eigenvalues using DTIFIT [75]. NODDI parameters (ICVF, ISOVF and ODI) were determined from the registered dMRI data using the NODDI Matlab toolbox (mig.cs.ucl.ac.uk; MatlabR2018b).

#### 2.6.7 dMRI processing: PSMD

Automatic calculation of peak width of skeletonized water diffusion parameters followed the procedure described by Baykara et al. (2016) [76] using their freely-available script (http://www.psmd-marker.com). Briefly, the dMRI data were processed using the standard Tract-based Spatial Statistics (TBSS; [77]) pipeline available in FSL with histogram analysis performed on the resulting white matter MD, λ_AX_, λ_RD_, FA, ICVF, ISOVF and ODI skeletons. First, all participants’ FA volumes were linearly and non-linearly registered to the standard space FMRIB 1 mm FA template. Second, a WM skeleton was created from the mean of all registered FA volumes. This was achieved by searching for maximum FA values in directions perpendicular to the local tract direction in the mean FA volume. An FA threshold of 0.2 was applied to the mean FA skeleton to exclude predominantly non-WM voxels. Third, MD, λ_AX_, λ_RD_, ICVF, ISOVF and ODI volumes were projected onto the mean FA skeleton and further thresholded at an FA value of 0.3 to reduce CSF partial volume contamination using the skeleton mask provided by [76]. Finally, PSMD, PSAD, PSRD, PSFA, PSICVF, PSISOVF and PSODI were calculated as the difference between the 95th and 5th percentiles of the voxel-based values within each subject’s DTI and NODDI skeletons.

#### 2.6.8 dMRI processing: Tractography

Quantitative tractography employs probabilistic neighbourhood tractography (PNT) as implement in TractoR, with the underlying connectivity data generated using FSL’s BedpostX/ProbTrackX tools run with a two-fiber model per voxel, 5000 probabilistic streamlines to reconstruct each tract with a fixed separation distance of 0.5 mm between successive points. Sixteen tracts of interest (Table 5) representing a wide range of projection, commissural and association fibers were identified in each subject using tract shape modeling in a 7 × 7 × 7 voxel neighbourhood. This allowed tract-specific mean values of DTI and NODDI biomarkers, weighted by connection probability, to be determined for each tract in every subject.

#### 2.6.9 MTI processing

Echoes for MTsat (On/Off/T1) volumes were summed together to increase signal-to-noise ratio (SNR) and the resulting MTsat-On and MTsat-T1 volumes were linearly registered to the MTsat-Off image with a rigid body transform (6 DOF). MTsat parametric maps were derived according to Helms [43] (https://doi.org/10.7488/ds/2965) and non-brain voxels were removed from the MTsat-Off image. MTR maps were also calculated from the MTsat-On and MTsat-Off images. This was followed by registration of the 3D T1W MPRAGE and tissue segmentations to the MTsat maps. Registration was done using the concatenated and inverted transformation matrix of a) linear registration of the MTsat-Off volume to the MTsat-T1 volume (7 DOF) and b) linear registration of the MTsat-T1 volume to the 3D T1W MPRAGE (12 DOF). Registered tissue segmentations were thresholded at 0.5, binarised and eroded by a sphere kernel of 1.4mm (with the exception of WML masks). The co-registered MTsat map and tissue segmentations were imported into MATLAB using spm_vol and spm_read_vols (SPM12, https://www.fil.ion.ucl.ac.uk/spm/software/spm12/). MTsat and MTR values within each tissue segmentation were then derived from respectively the masked MTsat and MTR map, thresholded at a range of 0 to 1 (MatlabR2018b).

#### 2.6.5 G-ratio

G-ratio calculation was performed by combining MTsat and dMRI measures. The MTsat map was registered to the first dMRI volume with the FSL FLIRT epi_reg script (FSL6.0.1), with registration to the bias-corrected 3D T1W MPRAGE as an intermediary step. The Myelin Volume Fraction (MVF) is calculated as: *MVF* = *MTsat* * *k*, where *k* was a constant derived by assuming a g-ratio of 0.7 in the splenium of the corpus callosum for two young, healthy control subjects, scanned twice [78]. NODDI and MTsat processing steps followed the patient pipeline, without a WML mask. The splenium mask was extracted from FreeSurfer segmentation of the T1-weighted MPRAGE structural image, registered to diffusion space.

The calibration factor was calculated as:

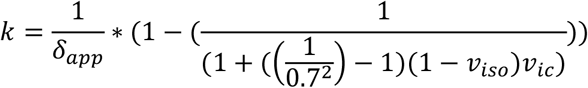

where *δ*_*app*_ is MTsat and *v*_*ic*_ and *v*_*iso*_ are the NODDI-derived intracellular volume fraction (i.e. neurite density index) and isotropic water diffusion. The mean k value across the splenium for each individual subject and each time-point was calculated and averaged across subjects and sessions.

The Axonal Volume Fraction (AVF) was calculated as:

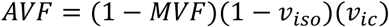

The aggregate g-ratio parametric maps were calculated voxel-by-voxel as [53]:

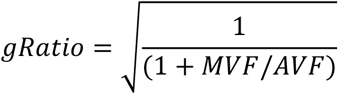

NAWM and WML segmentations were registered to the b0 diffusion volume with the transformation matrix from the FSL FLIRT epi_reg 3D T1W MPRAGE (FLS6.0.1) registration step, thresholded at 0.5 and binarised. G-ratio values within NAWM and WML segmentations were calculated.

#### 2.6.6. Relaxometry

Quantitative T1 maps were approximated from MT-off and MT-T1 images, using the same equations in Helms [43] (https://doi.org/10.7488/ds/2965). Processing steps followed the methodology for MTI. Quantitative T2 maps were generated using the two echo times from the dual echo sequence comprising T2W and PD, in protocol B (Table 3) using:

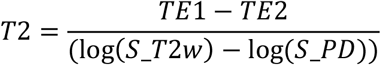

T2 maps were then linearly registered to the T1W image with a rigid body transformation (degrees of freedom = 6) and masked with tissue segmentations to derive tissue-specific quantitative T2 values.

### 2.7 Funding

FutureMS was funded by a grant from the Scottish Funding Council to Precision Medicine Scotland Innovation Centre (PMS-IC) and by Biogen Idec Ltd Insurance (R44346). The SWI sub-study was separately funded by Biogen Idec Ltd Insurance.

### 2.8 Study organization

FutureMS is a study hosted by PMS-IC and coordinated by the Anne Rowling Regenerative Neurology Clinic, University of Edinburgh. FutureMS participants were recruited and clinically assessed (including cognitive testing and neurological assessment) by trained medical personnel at the Anne Rowling Regenerative Neurology Clinic, University of Edinburgh; Glasgow Clinical Research Facility, Queen Elizabeth University Hospital; Aberdeen Clinical Research facility, Foresterhill Site, University of Aberdeen; Clinical Research Centre in Dundee, Ninewells Hospital; Raigmore Hospital in Inverness. MRI was performed at the Queen’s Medical Research Institute (QMRI) and the Royal Infirmary Edinburgh (RIE), University of Edinburgh; Clinical Research Facility Imaging Centre in Glasgow, Queen Elizabeth University Hospital; Aberdeen Biomedical Imaging Centre, Foresterhill Site, University of Aberdeen; Clinical Research Imaging Facility in Dundee, Ninewells Hospital. For participants recruited in Inverness, the MRI was performed at the site (Glasgow, Edinburgh, Dundee, and Aberdeen) most convenient for the participant. OCT imaging was performed at the Anne Rowling Regenerative Neurology Clinic, University of Edinburgh and Glasgow Clinical Research Facility, Queen Elizabeth University Hospital. FutureMS MRI data management and processing was performed at Edinburgh Imaging, Centre for Clinical Brain Sciences, University of Edinburgh. Genetic work streams were led by the University of California, San Francisco (UCSF), United States, and consisted of RNA sequencing and genotyping work performed at Edinburgh Genomics, University of Edinburgh and RNA analysis performed at UCSF, United States. Both MRI and genetic work streams were performed in close collaboration with the Anne Rowling Regenerative Neurology Clinic, University of Edinburgh.

### 2.9 Patient and public involvement

Principles of research transparency with study participants and shared research priority setting has been incorporated into the study design. Participants regularly receive written research updates participants are invited to join a voluntary network (Rowling Care) where they are kept up to date with research. They are also invited to in person research update presentations, which are planned to recommence in the near future having been delayed due to COVID-19 restrictions. Additionally, a sub-group of study participants meets regularly with the FutureMS research team and has been involved in setting study priorities and design.

## 3. SUMMARY

MRI allows the effects of MS on the brain to be probed non-invasively, provides potential specific biomarkers of underlying pathophysiology, and forms a core component of the overall FutureMS study. The current paper provides a detailed description of the FutureMS MRI protocol.

We have developed and implemented a comprehensive FutureMS MRI pathway that allows detailed capture of brain abnormalities in MS at both a microstructural and macrostructural level. This has involved development and testing of optimized and harmonized core protocols across MRI systems at multiple centres, resilient data transfer and QA procedures, and the largely automated data processing methods required for the high volume of imaging data generated from a large clinical cohort. Structured databases have been adapted and managed for large scale complex imaging datasets, which include both primary images and secondary processed data. Sub-studies have included additional ‘advanced’ MRI techniques; specifically, dMRI and MT targeted at reporting microstructural changes as quantitative biomarkers of demyelination and axonal degeneration characteristic in MS, and SWI as an indicator of chronic inflammation. We have applied widely-used and validated methods for processing conventional and advanced MRI data and adapted methodology from previous work for specific analyses, as required. These have allowed us to calculate conventional structural metrics such as WML volume and whole-brain and regional tissue atrophy, and generate masks corresponding to NAWM, GM and WML for calculation of microstructural measures within defined tissue types. Quantitative water diffusion and MT data allow advanced metrics such as NAWM and tract-specific myelin structure, myelin thickness and neurite density to be derived.

A possible limitation of the MRI component of FutureMS is the multicenter data acquisition and possible resulting variance in images between sites. However, multicenter MRI acquisition was required to access the MS population across Scotland and achieve adequate sample size and statistical power. A second limitation is the dual protocol implementation. Recruitment to FutureMS had started before there was an opportunity to develop and implement harmonized and optimized imaging protocols. These harmonized protocols are, however, important for a multicenter study to minimize variance in imaging measures across sites. To this end, the imaging protocol was updated after recruitment had already started, resulting in earlier participants having slightly different MRI examinations from those subsequently entering the study. Importantly, a pragmatic approach was therefore adopted to maximise imaging data consistency in which all participants received their baseline and follow-up imaging at the same site and on the same MRI system, using the same MRI protocol. Thirdly, MRI in Future MS is limited to brain, and will therefore not capture potentially important information on the role of spinal cord damage in disease severity and progression. However, possible inclusion of spinal cord imaging in a subset of participants at future follow-up visits is currently under consideration.

In conclusion, by integrating multimodal MRI, clinical, fluid biomarker and genetic data from a large population of RRMS patients in Scotland, FutureMS aims to develop predictive tools that will reduce uncertainty around disease progression and facilitate improved MS treatments.

## Data Availability

Data requests can be sent to the corresponding author. These will be handled on a case by case basis by the FutureMS steering committee and data custodians.

## ACKNOWLEDGEMENTS

We would like to thank Dr Pascal Sati for his help with implementing the SWI sequence in the FutureMS protocol. We would also like to thank other non-author contributors of the FutureMS consortium as follows: Chris Batchelor, Fraser Brown, Tracy Brunton, Yingdi Chen, Shuna Colville, Annette Cooper, Rachel Dakin, Liz Elliott, Peter Foley, David Hunt, Aidan Hutchison, Charlotte Jardine, Lucy Kesseler, Michaela Kleynhans, Jen MacFarlane, Bev MacLennan, Sarah-Jane Martin, Mary Monaghan, Scott Semple, Adam Scotson, Amy Stenson and Rosie Woodward. With special thanks to all FutureMS participants who have made this study possible.

FutureMS was hosted by Precision Medicine Scotland Innovation Centre (PMS-IC) and funded by a grant from the Scottish Funding Council to PMS-IC and Biogen Idec Ltd Insurance (R44346). Additional funding for authors came from the MS Society Edinburgh Centre for MS Research (grant reference 133; RM), NHS Lothian Research and Development Office (MJT), Chief Scientist Office – SPRINT MND/MS program (ENY) and the Row Fogo Charitable Trust (BROD.FID3668413; MVH). Additional funding for the Edinburgh university 3T MRI Research scanner in RIE is funded by the Wellcome Trust (104916/Z/14/Z), Dunhill Trust (R380R/1114), Edinburgh and Lothians Health Foundation (2012/17), Muir Maxwell Research Fund, Edinburgh Imaging, and University of Edinburgh.

## AUTHOR CONTRIBUTIONS

RM and SW wrote the manuscript with specific support from ENY, MEB, MVH, MJT, DM, KCNKK and ADW. RM and SW performed image data management, supported by DRG and DJ. MT devised and set up the MRI protocol across sites. RM, ENY, MEB, MVH, MT, NW and AK set up and performed image processing pipelines. DM, KCNKK and AW set up and performed qualitative MR assessment. CW performed overall FutureMS study coordination. PKAK performed clinical data management. PC and SC devised the original FutureMS study. ADW supervised the imaging part of FutureMS. All authors contributed to the final manuscript.

There are no competing interests to report.

